# Longitudinal determination of mRNA-vaccination induced strongly binding SARS-CoV-2 IgG antibodies in a cohort of Romanian healthcare workers

**DOI:** 10.1101/2021.03.17.21253751

**Authors:** Mónika Korodi, István Horváth, Kinga Rákosi, Zsuzsanna Jenei, Gabriella Hudák, Melinda Kákes, Katalin Dallos-Fejér, Enikő Simai, Orsolya Páll, Natalia Staver, Violeta Briciu, Mihaela Lupşe, Mirela Flonta, Ariana Almaş, Victoria Birlutiu, Claudia Daniela Lupu, Andreea Magdalena Ghibu, Dana Pianoschi, Livia-Maria Terza, Szilard N. Fejer

## Abstract

Mass vaccination against the disease caused by the novel coronavirus (COVID-19) was a crucial step in slowing the spread of SARS-CoV-2 in 2021. Even in the face of new variants, it still remains extremely important for reducing hospitalizations and COVID-19 deaths. Only limited data exists about the short- and long-term dynamics of humoral immune response. We present a longitudinal analysis of post-vaccination IgG levels in a cohort of 166 healthcare workers vaccinated with BNT162b2 with weekly follow-up until 35 days past the first dose and monthly follow-up up to 6 months post-vaccination. A subset of the patients continued with follow-up after 6 months and either received a booster dose or got infected during the Delta wave in Romania. Tests were carried out on 1697 samples using a CE-marked IgG ELISA assay developed in-house, containing S1 and N antigens of the wild type virus.

Participants infected with SARS-CoV-2 before vaccination mount a quick immune response, reaching peak IgG levels two weeks after the first dose, while IgG levels of previously uninfected participants mount gradually, increasing abruptly after the second dose. Overall higher IgG levels are maintained for the previously infected group 35-70 days after vaccination. The decrease of IgG levels is gradual, with lower overall values in the infection naïve cohort even 7-8 months after vaccination, compared to the previously infected cohort. Administration of a booster dose yielded higher average IgG antibody levels than post second dose in the infection naïve group and comparable levels in the previously infected group.

## Introduction

Vaccine-induced population immunity is an important step in the fight against the 2019 novel coronavirus (2019-nCoV)/severe acute respiratory syndrome coronavirus type 2 (SARS-CoV-2), and the coronavirus disease (COVID-19).^1^ Immunoglobulin G (IgG) is a good biomarker in blood for detecting long-term immune response due to infections.^2,3^ Infected individuals mount very different immune responses, and the antibody levels that can be measured post-infection have a large variation.^4–7^ Patients with moderate and severe symptoms have on average larger quantities of detectable antibodies,^8,9^ while those with mild symptoms or asymptomatic infections mount a weaker immune response, measurable by lower quantities of antibodies which often decrease below the detection threshold in a couple of months.^10^ However, the relationship between SARS-CoV-2 IgG antibody quantities and the level of protection is not yet established, especially in the light of the appearance of novel circulating variants, and is therefore subject to intense research. It is assumed that a subset of these antibodies, those capable of neutralizing the virus by interfering with cell attachment, has the biggest role for protective immunity, while other types of antibodies either contribute to protective immunity through other mechanisms (removal of infected cells),^11,12^ or cause long-term post-COVID complications.^13^ Experimental evidence for these mechanisms is scarce, and large observational follow-up studies are needed for determining any correlation between antibody levels and long-term protection from reinfection.^14^The matter is further complicated by vaccination-induced immunity. Currently approved mRNA vaccines in the European Union contain instructions for cells to synthesize the SARS-CoV-2 spike protein of the wild type virus, therefore the immune system will produce anti-SARS-CoV-2 spike antibodies,^15–17^ or contain the spike protein itself with an adjuvant.^18^ Several studies show a significant difference between post-vaccination immune response of previously infected individuals compared to uninfected vaccinated individuals,^19–21^ as the former group mounts a quick immune response within the first two weeks of the first vaccine dose, while the antibody titers of the non-infected group will be on average lower than those of the first group even 10 days after the second dose. However, much more data is needed to better understand the dynamics of post-vaccination antibody production in these two groups, especially in the context of waning immunity.

The BNT162b2 mRNA vaccine by BioNTech/Pfizer encodes the full-length transmembrane spike (S) glycoprotein, locked in its prefusion conformation by the substitution of two residues with proline.^22^ The available data shows that this vaccine had more than 90% efficacy in preventing COVID-19 in phase III clinical trials,^23–25^ while large-scale monitoring of more than 500,000 vaccinated individuals in Israel supported the result of phase III observations.^26^ However, vaccine efficacy against symptomatic infection was found to decrease over time, and this also depends on the variant in circulation. Latest data show significantly affected neutralization capacity of sera collected from fully vaccinated individuals against the Omicron (BA.1) variant.^27^

Unfortunately, vaccination coverage in Romania is very low, compared to other countries in Europe. Until 20 February 2022, only 43% of the population has been fully vaccinated, compared to the EU average of 72%. Romania experienced the biggest excess mortality among EU countries during the wave caused by the Delta variant (autumn 2021), due to the exceptionally low vaccination coverage among the high-risk population. Only limited data exists about post-vaccination kinetics among Romanian residents, although such data can help increase confidence of the population in the utility of vaccines.

Here we present results for longitudinal monitoring of strongly binding SARS-CoV-2 IgG antibodies in a group of 166 Romanian healthcare workers post-vaccination with the BNT162b2 vaccine, with a strict monitoring regimen that results in high-resolution per-patient data for the first four weeks post-vaccination. Antibodies produced against the SARS-CoV-2 nucleocapsid (N) and S1 domain of the spike protein were measured with separate in-house N and S1 enzyme-linked immunosorbent assays (ELISA) for a subset of these patients, and a CE IVD marked commercial version of these (SARS-CoV-2 IgG antibody detection kit, Proel Biotech), for the whole cohort. The commercial version is a combined N+S1 ELISA assay, optimized for low background signal. These assays were conceived to detect only IgG antibodies that have high affinity to the target antigens.

## Methods

### Study Design

This study is a prospective, multicenter cohort study aiming to monitor early and prolonged IgG production as a response to vaccination with the BNT162b2 COVID-19 vaccine. The study was carried out between January and December 2021 and consists of two periods. Period 1 lasted until two weeks past the second dose, with blood samples taken at one-week intervals to monitor the dynamics of early IgG production. We were aiming to collect four data points before the second dose, and at least one data point after the second dose from each participant in the first period. Period 2 shows the change in IgG levels 2-6 months after vaccination, with samples taken every four weeks, with the possibility of longer follow-up. A number of 48 patients opted for longer follow-up.

The population of the study was recruited among healthcare worker volunteers from 6 different medical centers (3 public hospitals and 3 private outpatient centers involved in diagnosis and treatment of COVID-19 patients). Written and informed consent was obtained from all participants. The study protocol was approved by the ethics committees of the three institutions involved in study design.

The study cohort consisted of 173 participants, from which 7 were excluded, as they did not meet the study criteria of at least 3 sample collections in Period 1, including at least one sample after 20 days of the first dose. 3 participants provided 4 samples, 1 participant 5 samples, 5 participants 6 samples, 4 participants 7 samples, 11 participants 8 samples, 28 participants 9 samples, 64 participants 10 samples, while 50 participants provided 11 or more samples. The total number of samples is 1697 for 166 participants.

In total, we present the results for 166 participants, 136 (82%) female and 30 (18%) male. The participants were aged between 22 and 71 years, the median age was 44 years. The age distribution of participants is the following: 38 between 15-34, 53 between 35-44, 49 between 45-54, and 26 people between 55-80. 25 participants had tested previously positive for SARS-CoV-2 infection with RT-PCR between August and December 2020, one participant had a confirmed infection in April 2020. Distinctly, 18 more participants had close contacts with COVID-19 patients and also had symptoms consistent with the disease but were not tested with RT-PCR. These healthcare workers had tested subsequently positive for SARS-CoV-2 IgG antibodies before getting vaccinated. The total number of confirmed previous SARS-CoV-2 infections is 64 (38.5%) within the study cohort.

### Development of the enzyme-linked immunosorbent (ELISA) assays

#### Preparation of SARS-CoV-2 antigen coated plates

The 96 well microplate (12×8 well strips on a single well holding plate frame, Greiner) was pre-activated at room temperature for 10 minutes with 50 µl/well Coating buffer 1x (Coating Buffer 10x concentrate, CANDOR Bioscience GmbH) - the 10x concentrate was diluted with deionized water to the appropriate concentration prior to use. After pre-activation, the content of the wells was completely aspirated. The microplates were coated with 50 µl antigen of 1 µg/ml concentration in Coating buffer, incubated at 4°C overnight. The antigen used for the S1 plates was the SARS-CoV-2 Spike S1 protein (His tag) from Sanyoubio, the one for the N plates was the SARS-CoV-2 nucleocapsid protein (His tag) from Sanyoubio. After incubation, the content of the wells was completely aspirated and 350 µl PlateBlock buffer (PlateBlock OEM, CANDOR) was added into each well of the microplate and incubated at room temperature for 2 hours, followed by aspirating the content of the wells. Antigens were preserved in their original conformation using 100 µl/well Liquid Plate Sealer (Liquid Plate Sealer OEM, CANDOR), incubated at room temperature for 90 minutes. After incubation, the content of the wells was completely aspirated. The coated plate was dried at 37°C by incubation for approximately 1 hour. The SARS-CoV-2 antigen coated plates were sealed with vacuum and stored at 4°C. The pre-activation, coating, blocking and sealing steps were carried out on an Opentrons OT-2 automated pipetting platform.

#### Assay procedure

Serum samples were diluted 1: 101 with Sample diluent (LowCross-Buffer OEM, CANDOR) and vortexed. 50 µl positive control (diluted human serum containing anti-SARS-CoV-2 IgG antibodies) and negative control (diluted human serum without anti-SARS-CoV-2 IgG antibodies) was added into the first two wells, and the diluted samples into consecutive wells of the antigen-coated plate and incubated at room temperature for 1 hour in the dark. After incubation, the wells were washed 5 times with 350 µl diluted Wash buffer (Washing Buffer PBS 10X OEM, CANDOR). 50 µl Enzyme Conjugate (Goat anti-Human IgG secondary antibody, Abcam, diluted 1:20.000 in LowCross HRP-Stab buffer, CANDOR) was added and incubated at room temperature for 15 minutes in the dark. After incubation, the washing step was repeated and 50 µl TMB Substrate solution (SeramunBlau automat fast, Seramun Diagnostica GmbH) was added into the wells, and another incubation step followed at room temperature for 20 minutes in the dark. After incubation, 25 µl Stop solution (0.25 mol/l sulfuric acid, Merck) was added into the wells and the optical density values at 450 and 620 nm (or 450 and 630 nm) were read on a microplate or microstrip reader (Adaltis Plab, StatFax 4700, StatFax 303+).

SARS-CoV-2 IgG binding antibodies were determined for all collected serum samples with a commercial version of the above-described ELISA assay, which includes both the S1 and nucleocapsid antigens coated on the microplate. The quantitative values were determined as a signal to cutoff ratio in arbitrary units (AU/ml), the cutoff was set to the optical density (OD) of the negative control plus 6x the standard deviation of OD values for pre-pandemic serum samples, (total 0.1 OD), measured during product validation in August 2020. The assay showed 100% specificity, determined on 88 pre-pandemic serum samples, and 100% sensitivity compared to a chemiluminescent assay containing an antigen different than the SARS-CoV-2 N or S1 protein, for 30 COVID-19 recovered patients 1-3 months after diagnosis. For the measurements reported here with the commercial version of the assay, samples were tested on the day of collection or frozen and thawed only one time. Subsequently, most collected samples during Period 1 were stored at -20°C and remeasured with the separate nucleocapsid and S1 assays in duplicates.

## Results

Figure 1 shows the dynamics of determined IgG values for each included participant in signal to cutoff type units (AU/ml) up to 70 days post-vaccination, divided into previously infected and uninfected groups. We observe two distinct patterns, with IgG values of the uninfected group well-separated from the other two groups within the first 21 days of vaccination. That separation is the most pronounced for data points acquired 12-16 days after the first vaccine dose. In line with other observations, the first vaccine dose for previously infected individuals acts as a booster dose, greatly increasing the detected antibody levels within a week. Only two, mildly symptomatic participants with previous positive PCR test results behaved significantly different from this group, showing a gradual mounting of immune response like the trends observed for previously uninfected individuals.

**Figure 1.**
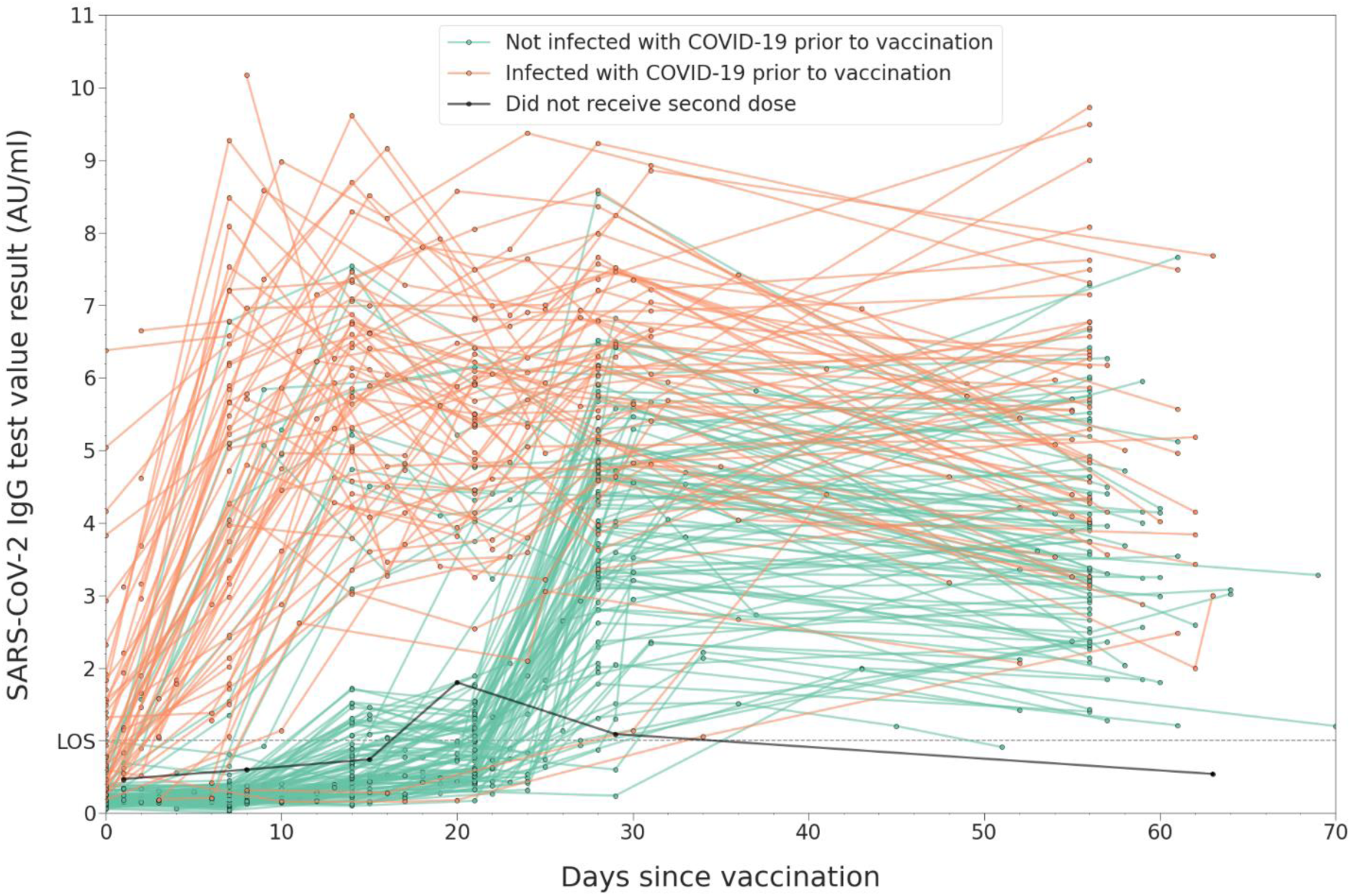
Dynamics of measured SARS-CoV-2 IgG values post-vaccination in the previously uninfected (infection naïve) and previously infected groups in the first 70 days after vaccination. Lines connect data points obtained longitudinally from the same patient. The limit of sensitivity (LOS) for the assay is 1 AU/ml.

After the second dose (day 21), an abrupt increase of IgG levels is observed for the previously uninfected group, while the change in antibody levels for the previously infected group is significantly more heterogenous, with a slight decrease observed for about the third of these participants. Nevertheless, at the end of Period 1 of the study, the average determined antibody levels for the previously infected group are significantly higher than those for the previously uninfected group.

Figure 2 shows the antibody dynamics in the infection naïve and previously infected groups. Antibody waning is first observed 36-65 days post vaccination. The overall slight decrease of IgG levels becomes more evident in the previously uninfected group 66-95 days post-vaccination. One female participant from the previously uninfected group did not receive the booster dose, and her IgG level measured 63 days after vaccination got below the limit of sensitivity, in the range of the assay that is interpreted as negative. That participant was subsequently infected during the spring wave in Romania, caused by the Alpha variant and developed detectable antibodies after infection. Apart from this participant, we observe a single, previously uninfected weak responder to the vaccine, with peak IgG levels of only 1.51 AU/ml after the second dose, decreasing to 0.91 after 51 days post-vaccination (>0.9 AU/ml, within the ‘inconclusive’ range of the assay).

**Figure 2.**
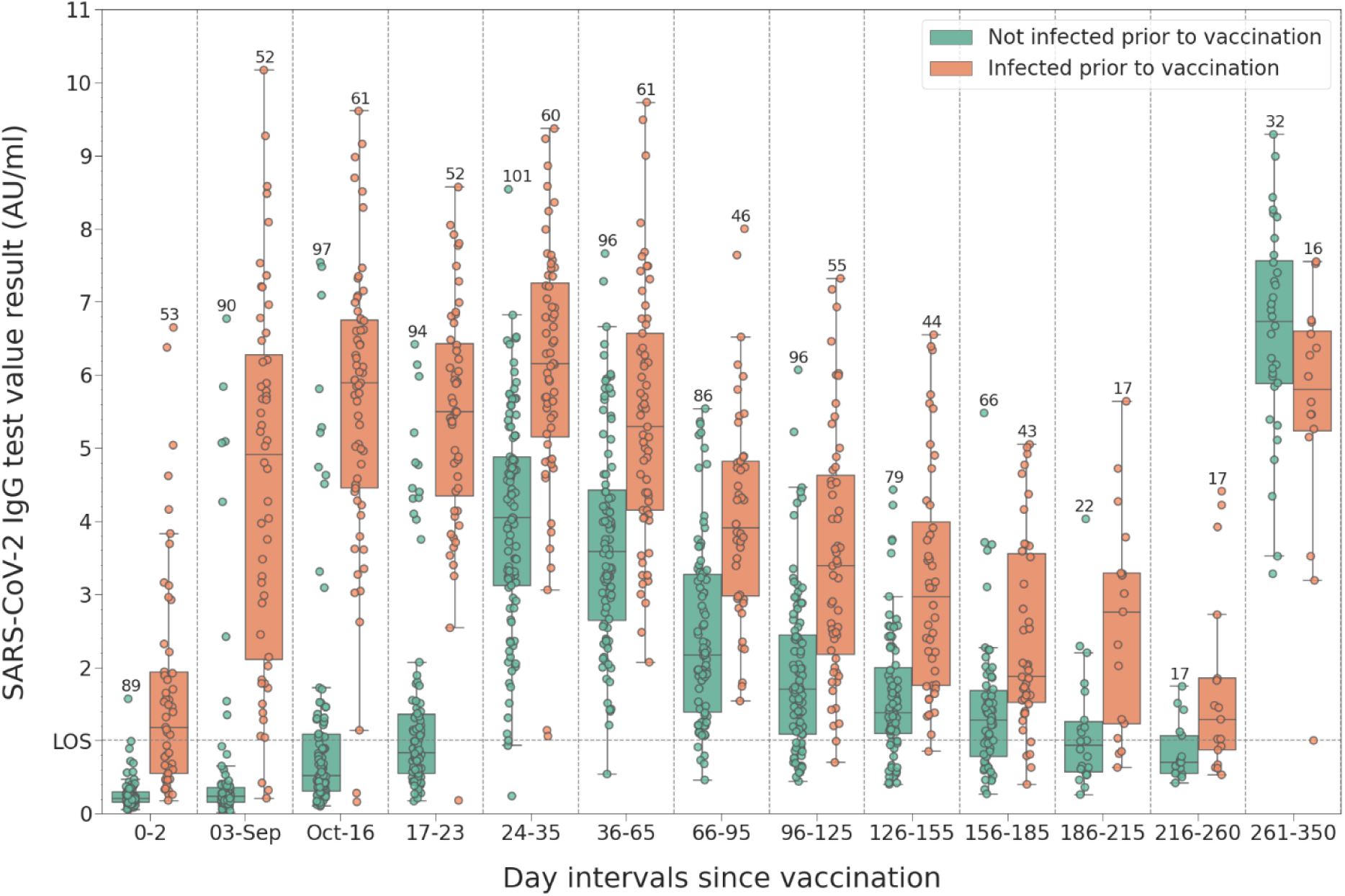
Distribution of measured SARS-CoV-2 IgG binding antibody levels at different time intervals post-vaccination with the first dose: baseline (0-2 days) and weekly intervals until the administration of the second dose. If multiple measurements were done for a participant within these intervals, only the largest value is shown. The last interval shows the immune response of participants who opted for long-term follow-up. Most of these participants received the booster dose in this interval, and 8 were infected after two doses. The number of data points in each group is shown above the highest value.

The influence of booster doses or infection during the study period is shown in Figure 3. The antibody values are shown for those participants who opted for long-term follow-up and either received a booster dose or got infected during the study duration. This latter cohort contains two participants infected during the spring wave (Alpha variant), and 8 participants infected during the autumn wave (Delta variant), as confirmed by positive RT-PCR. Although the sample size is small, it can be seen that the booster dose significantly increases peak IgG levels in the infection naïve cohort, way above peak levels observed after two doses. However, in the previously infected group (n=10), post-booster peak IgG levels increase only to values similar to those observed after two doses. The biggest observed increase in peak IgG levels is that for the group that opted for long-term follow-up and got infected more than 8 months after vaccination (last interval from Figure 3, n=8).

**Figure 3.**
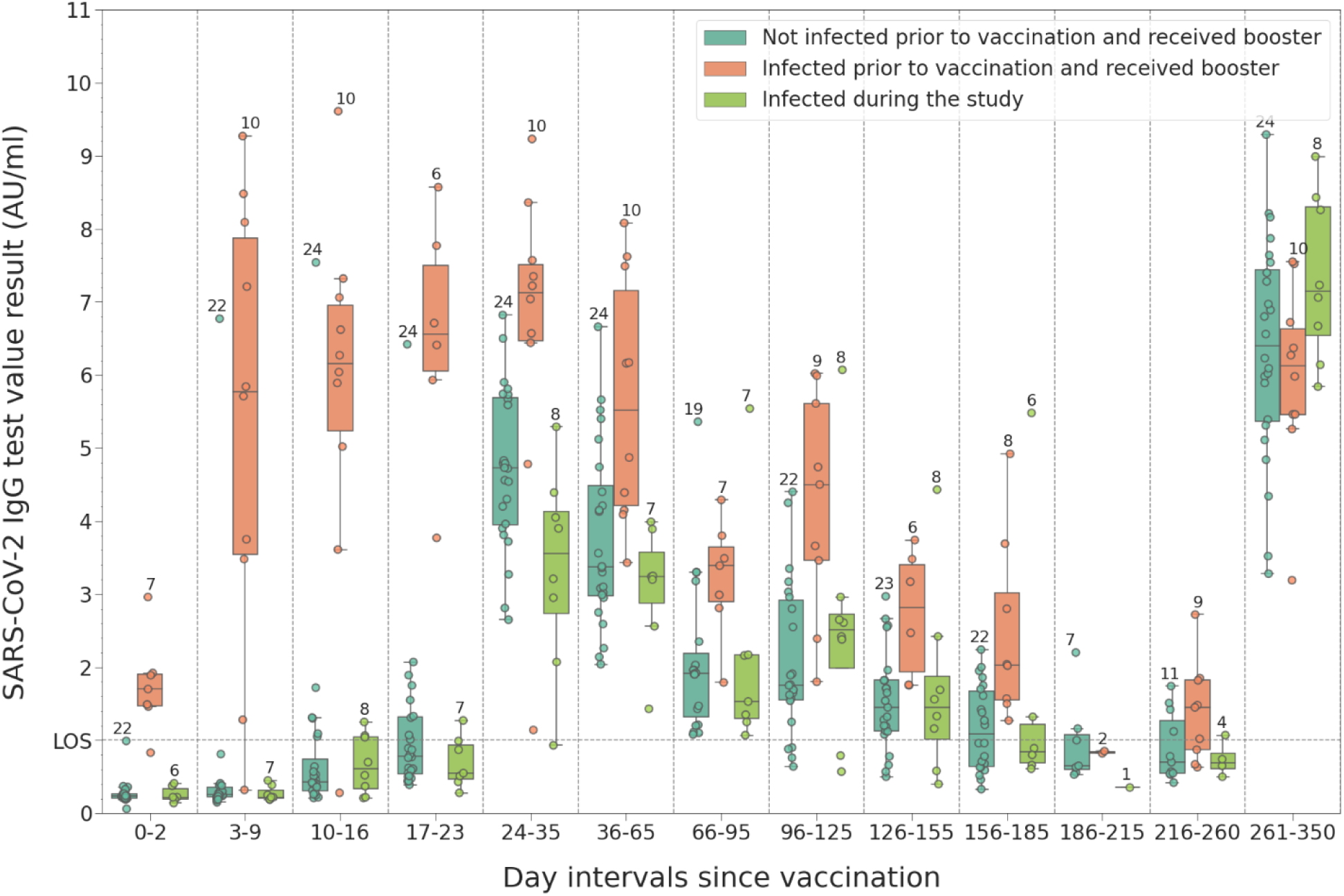
SARS-CoV-2 IgG binding antibody dynamics for a subgroup of the participants, those who either received a booster dose 9-10 months after vaccination or were infected during the duration of the study.

The first two groups in Figure 4 are the same as those in Figure 3 (participants that received the booster dose). The increase in antibody levels for the third group approx. 9 months post-vaccination (infection naïve participants who did not receive booster) is due to infection during the Delta wave, as confirmed by positive RT-PCR. As it can be seen from the dynamics of the last group (previously infected participants that did not receive the booster dose), it is likely that most were reinfected during the Delta wave, shown by the increase in antibody values in the last period of the study.

**Figure 4.**
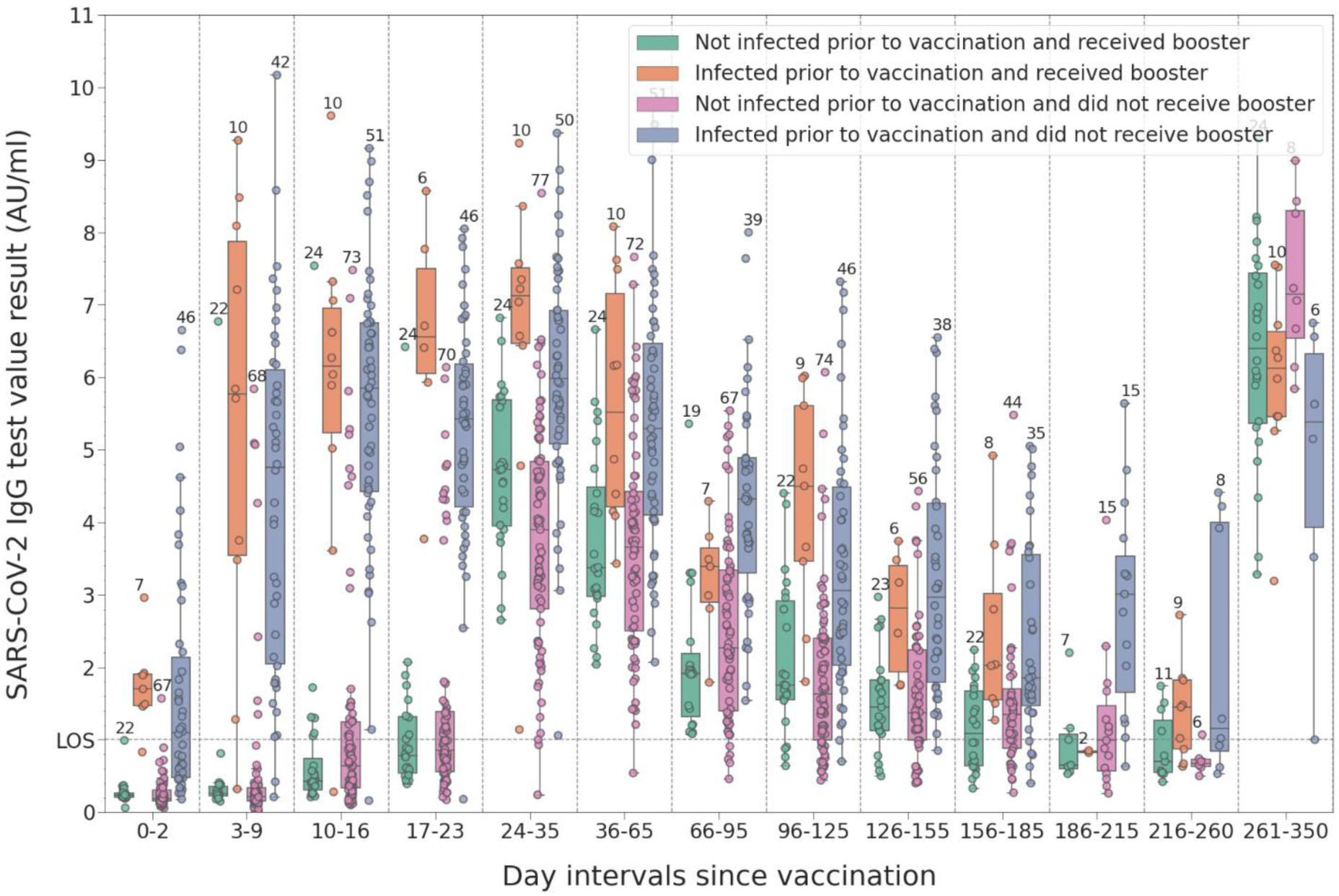
Distribution of measured SARS-CoV-2 IgG binding antibody levels at different time intervals post-vaccination with the first dose: baseline (0-2 days) and weekly intervals until the administration of the second dose. Participants were grouped according to their pre-vaccination infection status and whether they received a booster during the last period or not.

As the commercial version of our ELISA assay detects both anti-N and anti-S1 IgG antibodies, we remeasured the samples with separate anti-N and anti-S1 in-house assays to observe the dynamics of the two antibody types individually for Period 1 of the study. Supplementary Figure 1 shows the obtained per-participant plots and the corresponding box plots for the same intervals of Period 1 as in Figure 2. As expected, vaccination does not have an effect to observed anti-N levels, the longitudinal increase is due to vaccination-induced anti-S1 antibody production, with similar dynamics as that measured with the commercial assay, with the exception of the baseline (0-2 days) in the previously infected group, where we observe much lower quantities of strongly binding anti-S1 antibodies than anti-N antibodies. The difference between *baseline* antibody levels of the two groups, measured with the commercial version of the assay, is therefore mainly due to the presence of anti-N antibodies in the previously infected group.

Supplementary Figure 3 shows the change in measured SARS-CoV-2 IgG values grouped according to the gender of participants and their previous infection status. We observe no significant difference between the immune response of male and female participants, the resulting deviations among the previously infected group can be due to the small sample size for male participants. However, the age distribution shows different behavior based on post-vaccination immune response among the previously infected and uninfected groups, as shown in Supplementary Figure 4. Among the uninfected group, antibody levels increase fastest within the young cohort (age groups 22-34 and 35-44). Among the previously infected group, the participants aged over 55 years have larger baseline antibody levels and their post-vaccine immune response is significant enough to keep antibody levels higher than those measured for the groups aged 22-54 at the end of Period 1 and even 36-70 days post-vaccination, although the number of data points is much smaller for this period. As post-infection antibody levels are correlated with disease severity, which is also correlated with age, it is possible that the observed post-vaccination humoral immune response of previously infected older participants is stronger due to the larger baseline antibody levels mounted as a response to more severe symptoms compared to the younger population.

## Limitations

Our study has several limitations. The number of participants can be considered small, compared to much larger studies in this field, for example that of the COG-UK consortium. However, we are not aware of other similar studies that collected such a high resolution longitudinal data for the first 5 weeks post-vaccination from participants. Another limitation is that we detect strongly binding antibodies instead of neutralizing antibodies, although these two are likely in a strong correlation. We did not attempt to intercompare the measured antibody values with more mainstream methods of large diagnostic manufacturers, but we did calibrate our CE-marked assay against the first international WHO standard for SARS-CoV-2 antibodies (First WHO International Standard for anti-SARS-CoV-2 immunoglobulin (human), NIBSC code: 20/136. National Institute for Biological Standards and Control, 2020). As the standard was widely accessible only after the start of our study, we could not specify our values in international units. However, we provide calibration curves for some of the lots used in the study in Supporting Information to aid for assessing the reproducibility of our results.

## Discussion

Our results show a robust humoral immune response that mounts gradually after the first BNT162b2 vaccine dose for the group previously uninfected with SARS-CoV-2, and a much stronger immune response within 7-14 days after the first dose for the previously infected group, in line with recent data from other cohorts of healthcare workers.^19^ For the previously infected group, the second dose slightly elevates the SARS-CoV-2 anti-S1 antibody levels further, and we do not see the levels of the two groups converging even after 7 months post-vaccination. Our long-term follow-up results, albeit on a small sample, suggest that infection (with the Delta variant) *after* vaccination of previously infection naïve individuals drives a more robust humoral immune response than infection *before* vaccination followed by a booster dose.

The kinetics data obtained within the first three weeks for the previously infected group suggests that a SARS-CoV-2 infection followed by a single BNT162b2 vaccine dose produces larger quantities of strongly binding anti-spike 1 IgG antibodies than two BNT162b2 doses three weeks apart in previously uninfected individuals. In order to maximize antibody levels produced by uninfected vaccinated individuals, it was proven before that delaying the second dose by a few weeks boosts the overall immune response relative to the 3-week dosing regimen. However, delaying the second dose might reduce protection for immunocompromised individuals,^28,29^ with an unknown increase of infection risk even among a healthy population, as the minimal amount (and quality) of circulating antibodies necessary for protective immunity against SARS-CoV-2 infection is unlikely to be determined yet, especially in light of novel circulating variants.^27,30^ Given the shortage of available vaccines, especially in developing countries, and the urgent need to confer long-lasting protection from severe forms of the disease, antibody-based approaches might be needed in the future to optimize vaccine distribution, dosing regimens, and the maximal length of acquired immunity against COVID-19 among various population groups.

## Supporting information

Supplementary discussion, figures and tables

## Data Availability

Anonymized data for each sample (age, gender, day elapsed since the first vaccine dose) and ELISA measurements for SARS-CoV-2 IgG antibodies are available in a tabular form.

https://szilard.ro/files/covid_antibody_study_complete.csv

## Notes

### Competing Interest Statement

SNF is the CEO of a startup of Pro-Vitam Ltd, and head of the group that developed the combined S1+N ELISA assay, commercialized by the startup.

### Funding Statement

Research costs were supported by each participating institution. No other funding to disclose.

### Author Declarations

The study protocol was approved by the ethics committees of the Hospital for Infectious Diseases Cluj-Napoca (approval no. 756/13.01.2021), of the Infectious Diseases Clinic, Academic Emergency Hospital Sibiu (2238/29.01.2021), and of Pro-Vitam Ltd (1711/05.01.2021).

### Summary of Updates

Longitudinal data was collected up to a maximum of 11 months post-vaccination for a subset of participants. Data analysis now contains 2.5 times more data points than the original version, the total number of participants was increased at 166. Our long-term follow-up results, albeit on a small sample, suggest that infection (with the Delta variant) AFTER vaccination of previously infection naive individuals drives a more robust humoral immune response than infection BEFORE vaccination followed by a booster dose.

