## Supplementary discussion, figures and tables for "Longitudinal determination of mRNA-vaccination induced strongly binding SARS-CoV-2 IgG antibodies in a cohort of Romanian healthcare workers"

### Supplementary Material for the paper "Longitudinal determination of mRNA-vaccination induced strongly binding SARS-CoV-2 IgG antibodies in a cohort of Romanian healthcare workers"

Mónika Korodi[1,2], István Horváth[1,2], Kinga Rákosi[1,3], Zsuzsanna Jenei[1], Gabriella Hudák[1], Melinda Kákes[4], Katalin Dallos-Fejér[1], Enikő Simai[4,5], Orsolya Páll[1,2], Natalia Staver[4], Violeta Briciu[6,7], Mihaela Lupșe[6,7], Mirela Flonta[6], Ariana Almaș[6], Victoria Birlutiu[8,9], Claudia Daniela Lupu[8], Andreea Magdalena Ghibu[8], Dana Pianoschi[6], Livia-Maria Terza[1], Szilard N. Fejer[1,2,10*]

1 Pro-Vitam Ltd., Muncitorilor street 16, Sfantu Gheorghe, Covasna, Romania, 520032
2 Department of Chemistry, University of Pécs, Ifjúság street 6, Pécs, Hungary, 7624
3 Faculty of Technical and Human Sciences, Sapientia Hungarian University of Transylvania, Corunca 1c, Targu Mures, 540485
4 Promedical Center Ltd., Caisului street 16, Cluj-Napoca, Cluj, Romania, 400487
5 Benedek Geza Hospital for Rehabilitation of Cardiovascular Disease, Mihai Eminescu street 160, Covasna, Covasna, Romania, 525200
6 Clinical Hospital of Infectious Diseases, Iuliu Moldovan street 23, Cluj-Napoca, Cluj, Romania, 400348
7 Department of Infectious Diseases, University of Medicine and Pharmacy Iuliu Hatieganu, Iuliu Moldovan street 23, Cluj-Napoca, Cluj, Romania, 400348
8 Faculty of Medicine, Lucian Blaga University of Sibiu, 2A Lucian Blaga street, Sibiu, Sibiu, Romania, 550169
9 Infectious Diseases Clinic, Academic Emergency Hospital Sibiu, 2-4 Corneliu Coposu Bld. , Sibiu, Sibiu, Romania, 550245
10 Proel Biotech Ltd., Muncitorilor street 16, Sfantu Gheorghe, Covasna, Romania, 520032

#### Supplementary discussion

To verify whether the significant difference between antibody levels measured with the N+S1 combined commercial assay at the start of Period 2 (36-65 days after vaccination) is due to high maintained anti-N IgG values in the previously infected group, we remeasured the the majority of samples collected at the baseline and those collected for Period 2, with the ingredients of the commercial assay, except that the plates were coated separately with N and S1 proteins according to the procedure described in the Methods section. We observe an overall decrease in anti-N IgG levels compared to the baseline in the previously infected group (Supplementary Figure 2, left panel), while the difference of anti-S1 IgG levels between the uninfected and previously infected groups remains significant (p<0.0107, Supplementary Figure 2, right panel).

The reduced sensitivity of the assay for anti-S1 IgG in the previously infected group, compared to that of the anti-N assay, cannot be explained by the difference in antigen size alone, since for a hypothetical case of a patient producing anti-N and anti-S1 antibodies of equal concentration and affinities, the size difference would decrease the signal of anti-S1 antibodies by 40% at most, corresponding to the 1 µg/ml coating solution containing less S1 proteins (1.06 fmol/well N protein *versus* 0.65 fmol/well S1 protein). It is likely therefore that patients produce anti-S1 antibodies that bind with affinities below the threshold our assay can detect. However, high affinity anti-S1 antibodies should have an important role in ensuring protection from infection, since a subset of these would be the most potent in preventing cell attachment.

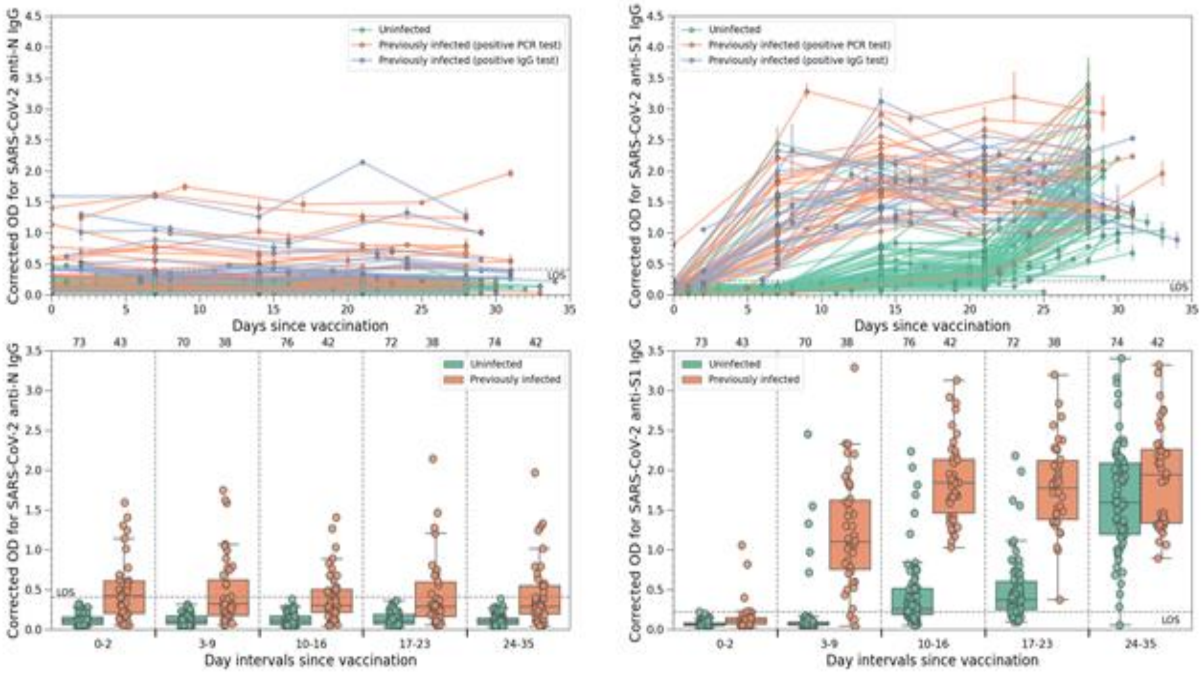


**Figure S1**. Dynamics of measured SARS-CoV-2 anti-N and anti-S1 IgG levels among the study participants over time for Period 1 of the study (0-35 days post-vaccination). The numbers on top of each box plot represent the number of samples within each group. Top column: anti-N IgG levels in optical density units (ODU), right column: anti-S1 IgG levels on the same scale.

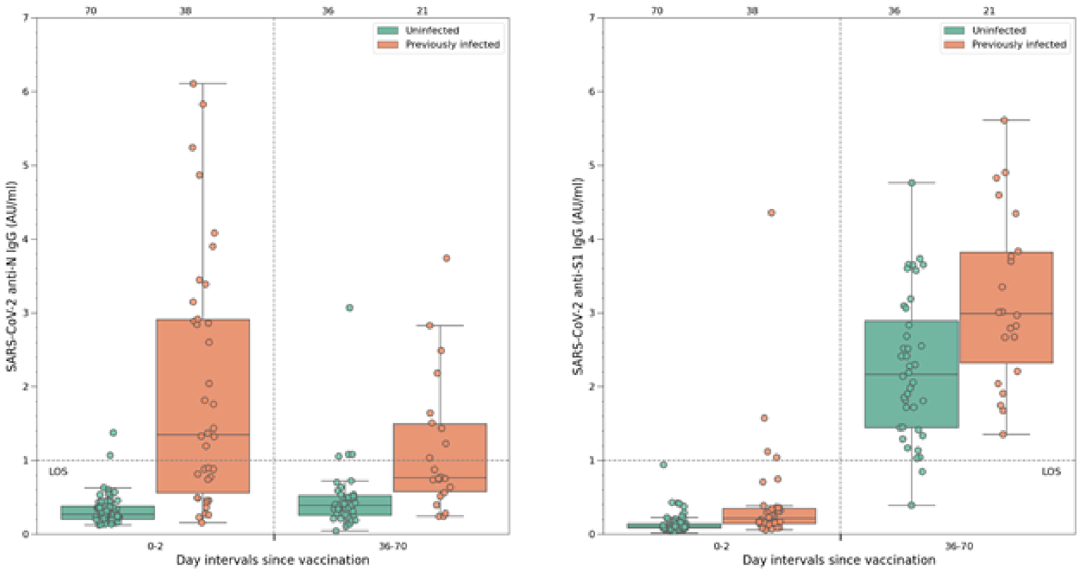


**Figure S2.** SARS-CoV-2 IgG levels for the uninfected and previously infected groups at the start of vaccination and 36-70 days after the first dose. Left panel: anti-N, right panel: anti-S1 IgG antibodies determined on nucleocapsid-only and S1-only plates. Apart from the concentration of coated antigens, for this measurement we used the same assay ingredients and protocol as that in the CE-marked commercial version. If multiple measurements were done for a participant within these intervals, only the largest value is shown.

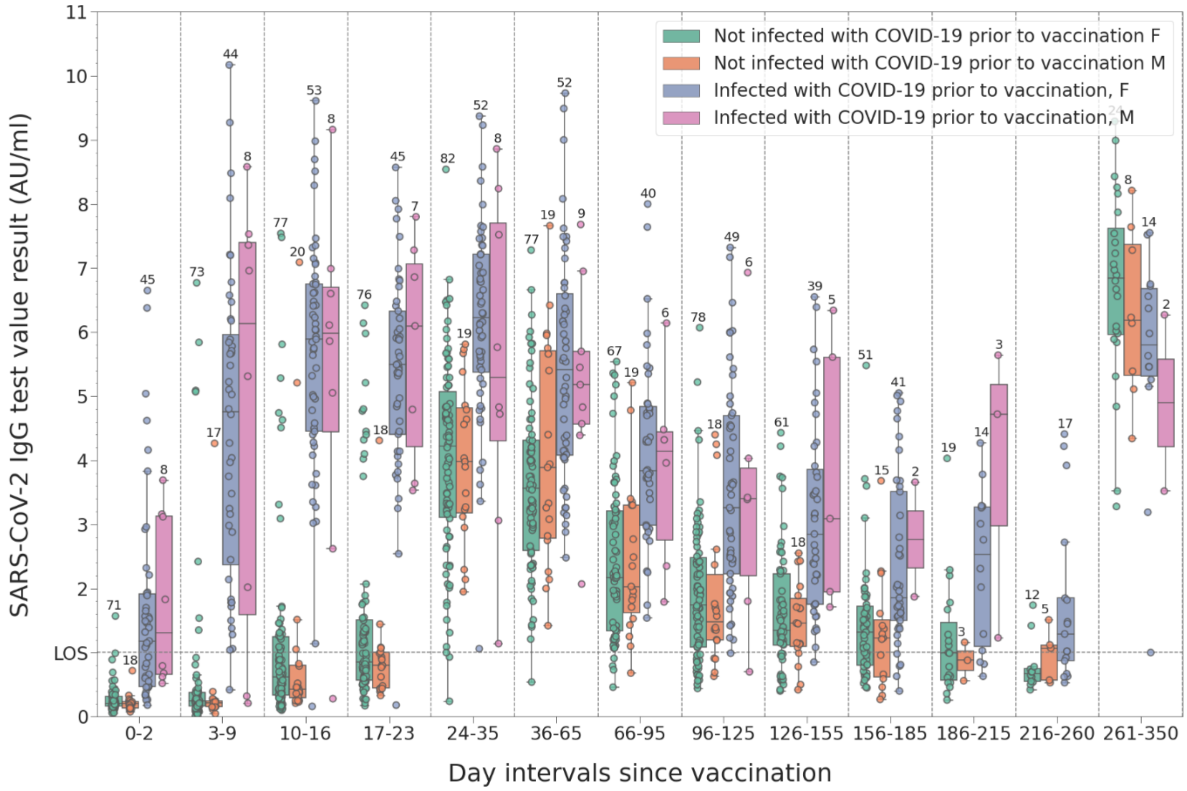


**Figure S3.** Longitudinal change of determined SARS-CoV-2 IgG values based on the gender and previous infection status of the participants.

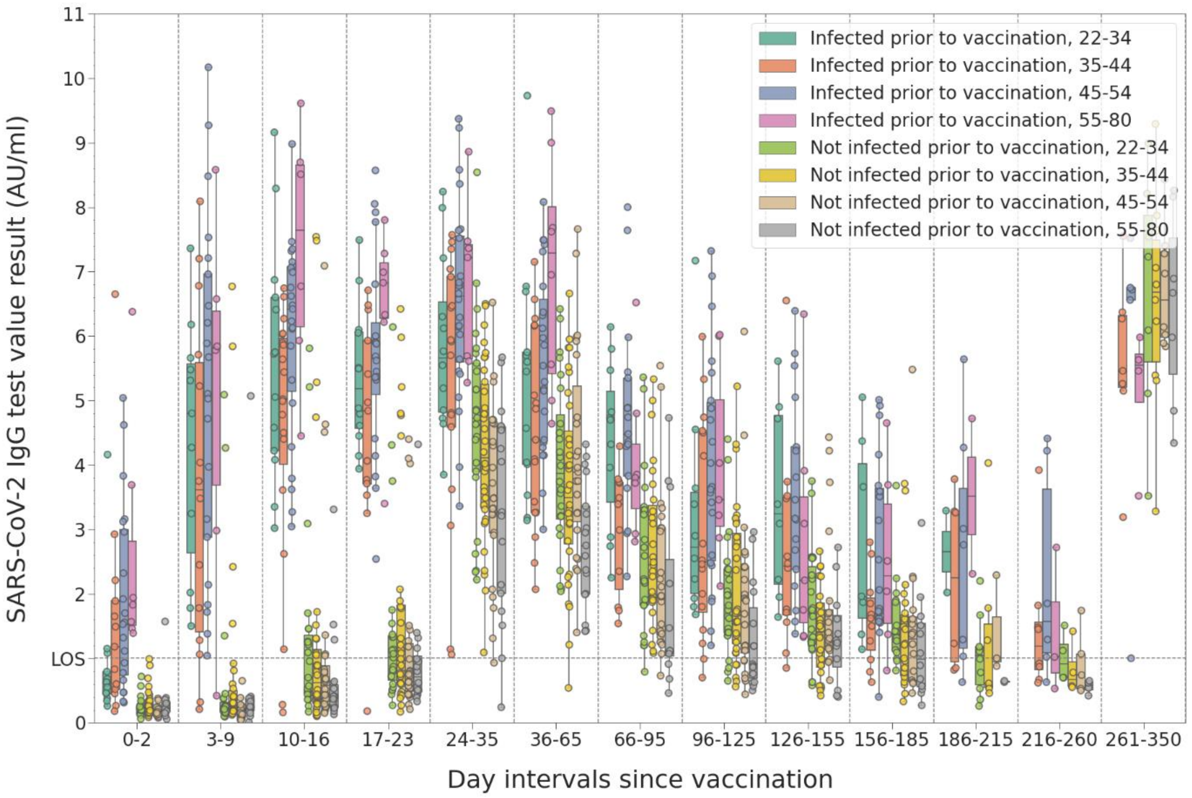


**Figure S4.** Longitudinal change of determined SARS-CoV-2 IgG values grouped according to the age group and previous infection status of the participants.

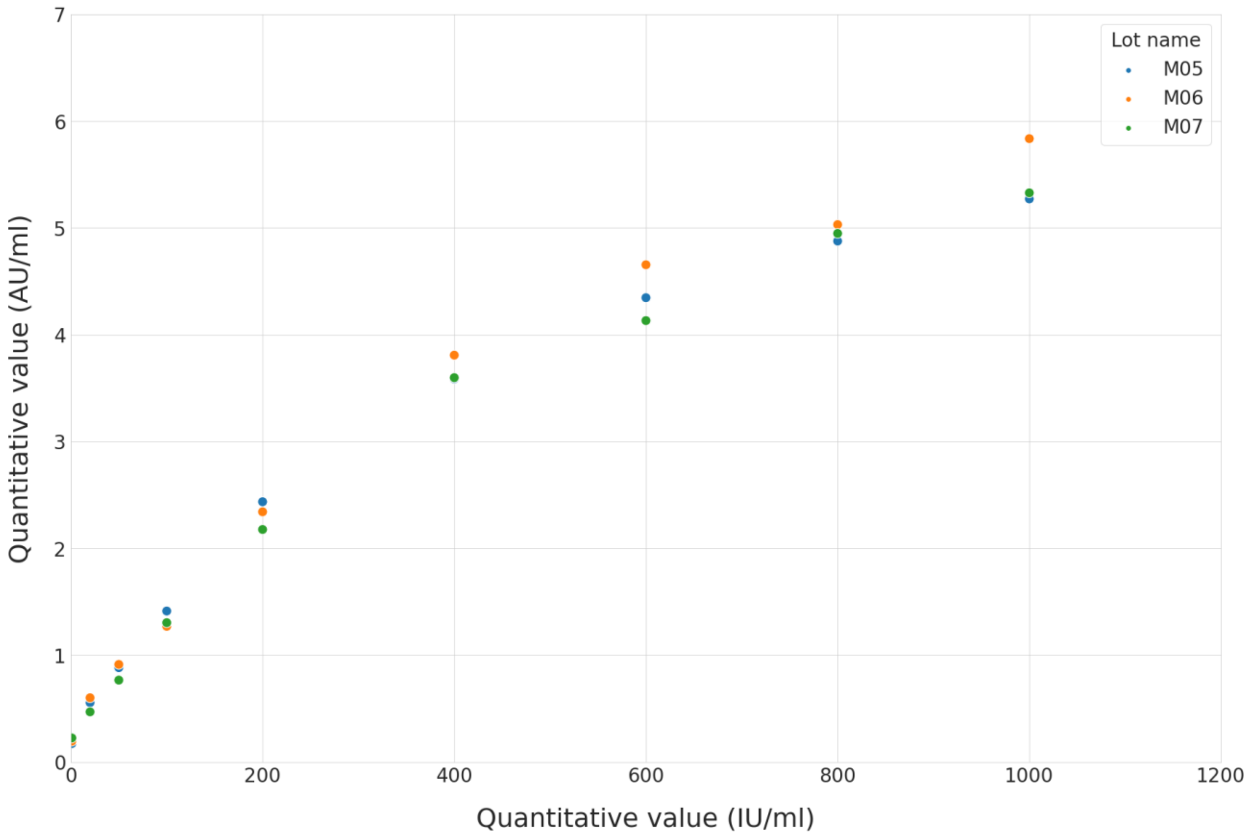


**Figure S5.** Correlation between SARS-CoV-2 IgG antibody values measured in IU/ml (using the first WHO standard) and AU/ml of the CE-marked version of the assay used in this study.

Supplementary Tables 1 and 2 show the distribution of study participants according to their gender and ages, compared to that of the available statistical data from Romanian healthcare workers.

**Table S1.** Demographics of the study participants based on their genders and three groups: all patients, the ones with an infection recorded prior to vaccination and the ones with no recorded infection prior to the vaccination. As the p-values show, all groups can be considered as representative of the population of healthcare workers in Romania.

| Group | Sample (Study) | Population (Healthcare statistics) | p-value |
|---|---|---|---|
| **Gender (F) all** | 81.93% | 79.30% | 0.517 |
| **Gender (F) with prior infection** | 84.38% | 79.30% | 0.21 |
| **Gender (F) with no known prior infection** | 80.39% | 79.30% | 0.787 |

**Table S2.** Demographics of the study participants based on their ages and three groups: all patients, the ones with an infection recorded prior to vaccination and the ones with no recorded infection prior to the vaccination. As the p-values show all groups can be considered as representative of the population of healthcare workers in Romania.

| Group \ Ages | 15 - 34 | 35 - 44 | 45 - 54 | 55 - 80 | p-value |
|---|---|---|---|---|---|
| **Population** | 20.10% | 37.00% | 27.00% | 15.90% | N/A |
| **All participants** | 20.48% | 23.49% | 22.89% | 13.25% | 0.111 |
| **Prior infection** | 18.75% | 25.00% | 28.12% | 6.25% | 0.02 |
| **No recorded prior infection** | 21.57% | 22.55% | 19.61% | 17.65% | 0.047 |

**Table S3.** P-values between IgG levels measured for infection naïve and previously infected groups, as shown in Figure 2.

| Day interval | p-value (between previously infected - not infected groups) |
|---|---|
| 0-2 | <0.001 |
| 3-9 | <0.001 |
| 10-16 | <0.001 |
| 17-23 | <0.001 |
| 24-35 | <0.001 |
| 36-65 | <0.001 |
| 66-95 | <0.001 |
| 96-125 | <0.001 |
| 126-155 | <0.001 |
| 156-185 | <0.001 |
| 186-215 | <0.001 |
| 216-260 | 0.006 |
| 261-350 | 0.022 |
